# Efficacy of acupuncture treatment for generalized anxiety disorder: a protocol for systematic review and meta-analysis

**DOI:** 10.1101/2025.04.26.25326498

**Authors:** Cláudio Calixto Carlos da Silva, Marcos André de Matos, Sandro Rogério Rodrigues Batista, Rodolfo Rêgo Deusdará Rodrigues, Neysa Aparecida Tinoco Regattieri

## Abstract

**Background:** Generalized Anxiety Disorder (GAD) is a prevalent mental health condition, characterized by excessive anxiety and worry that interferes with daily life. Many individuals with GAD do not receive timely and appropriate treatment, which can lead to chronic symptoms and a diminished quality of life. In addition to its psychological burden, GAD is associated with multiple physical symptoms, including tremors, muscle twitches, pain, nervousness, excessive sweating, diarrhea, palpitations and shortness of breath. As a result, it represents a significant public health challenge. Acupuncture has gained attention as a potential therapeutic approach for GAD. However, clinical research on its efficacy remains inconclusive. This systematic review aims to compile and critically evaluate the available evidence on the effectiveness of acupuncture in the treatment of GAD.

**Methods and Analysis:** A comprehensive search will be conducted across eight electronic databases: EMBASE, LILACS, PubMed/MEDLINE, SCIELO, SCOPUS, CINAHL, Web of Science, and the Cochrane Library. Additionally, all relevant studies from the gray literature in English, Portuguese, or Spanish will be considered. The primary outcome measure will focus on anxiety improvements, assessed using validated questionnaires. Risk of bias assessment, data synthesis, and subgroup analysis will be performed using Review Manager 8.1.1.

**Discussion:** This systematic review aims to evaluate the efficacy of acupuncture for GAD using Western databases. While limitations include the exclusion of Chinese databases and potential study heterogeneity, this review seeks to expand the understanding of acupuncture as a therapeutic option for GAD.

**PROSPERO registration number:** PROSPERO CRD42025622599

## Introduction

### Overview of the Condition

Generalized Anxiety Disorder (GAD) is characterized by excessive anxiety and worry related to various events or activities, lasting more than six months. The intensity, frequency or duration of these worries is often disproportionate to the likelihood or actual impact of the anticipated event, significantly compromising psychosocial functioning^1^. An estimated 264 million adults worldwide suffer from anxiety disorders, with a higher prevalence in women and adults between 45 and 55 years of age^2^. The pathophysiology of GAD involves multiple neurotransmitter systems, including serotonin, norepinephrine, glutamate, and cholecystokinin, which contribute to the dysregulation of arousal and sleep, often resulting in restless and unsatisfying sleep^3^.

### Therapeutic Approach and Mechanism of Action

The treatment of generalized anxiety disorder (GAD) should be individualized, considering factors such as the severity of the condition, the presence of comorbidities, and the patient’s treatment history. Therapeutic approaches include psychoactive medications, such as paroxetine, escitalopram, and sertraline, as well as psychotherapy, physical exercise, and complementary interventions^4^. However, many patients struggle to achieve satisfactory outcomes with pharmacotherapy alone^5^. As a result, an ideal treatment for GAD has yet to be developed, highlighting the need for more comprehensive and personalized strategies.

Acupuncture, a practice of traditional Chinese medicine, involves the insertion of fine needles into the skin at specific points and has been widely used in the management of mental health disorders^6^. Two recent systematic reviews with meta-analyses have demonstrated the positive effects of acupuncture on generalized anxiety disorder (GAD), highlighting its good tolerability and low incidence of side effects^7,8^. Acupuncture modulates key neurobiological mechanisms involved in anxiety regulation by influencing multiple pathways, such as the opioidergic, endocannabinoid, serotonergic, and hypothalamic-pituitary-adrenal axis. These effects help restore balance in the central and autonomic nervous systems, supporting acupuncture as a potential therapeutic intervention for Generalized Anxiety Disorder (GAD)^9-12^.

### The importance of conducting this review

Despite advancements in research, there remain gaps in systematic reviews based on ongoing studies conducted in the West. As previous systematic reviews have primarily focused on studies conducted in China, which are scarcely available in Western databases^7,8^. To address this, the present systematic review will focus exclusively on Western databases, aiming to investigate and describe the efficacy of acupuncture as a treatment for Generalized Anxiety Disorder (GAD). This review seeks to provide a comprehensive analysis of current evidence, contributing to a deeper understanding of acupuncture’s role in managing GAD within Western medical contexts.

## Materials and Methods

### Protocol Registration and Reporting Guidelines

This protocol is registered in the International Prospective Register of Systematic Reviews (PROSPERO) under the registration number CRD42025622599. The study follows the Preferred Reporting Items for Systematic Reviews and Meta-Analyses (PRISMA) guidelines. The PRISMA-P checklist for this protocol is provided as S1 Appendix.

### Eligibility criteria

The following inclusion and exclusion criteria will be applied. The review question is: Is acupuncture and its related techniques compared to usual treatment effective therapies for treating GAD in adults? The PICOS, which consists of participants/populations, interventions, comparator group, outcomes and study designs, is often used to refine the review question. The PICOS of this protocol is as follows.

### Types of participants

Participants with a confirmed diagnosis of GAD based on internationally recognized diagnostic criteria by the International Classification of Diseases (ICD-11)^13^ or Diagnostic and Statistical Manual of Mental Disorders-5 (DSM-V)^1^, adults over 18 years of age, both sexes.

### Types of interventions

Studies describing an observation using acupuncture and its related techniques, including dry needling, manual acupuncture, craniopuncture, auriculotherapy, moxibustion, laser acupuncture and/or electroacupuncture. There are no limitations on techniques used, selection of acupuncture points, types of needles, duration of treatment, or treatment protocols. For clinical trials, those who describe an intervention using any of the acupuncture modalities and compare it with a control group will be considered. Treatment protocols of any duration using any of the techniques mentioned above will be accepted.

### Types of comparator groups

The control group should not receive interventions using acupuncture and its modalities, but may have received an active intervention, that is, usual pharmacological medications and psychotherapy, or no treatment such as waiting list and placebo controls: such as sham acupuncture, placebo medications and simulated interventions. Sham acupuncture includes electrical stimulation without needling, needling without stimulation, needling using non-penetrating needles, or needling at points other than those selected in the intervention group.

### Types of Outcome Measures

Studies that evaluate the effectiveness of interventions using acupuncture and/or its related techniques in individuals with GAD will be included.

#### Primary outcomes

1. Assessment of anxiety using valid and reliable instruments: includes the Hamilton Anxiety Scale (HAM-A)^14^, Generalized Anxiety Disorder Scale (GAD-7)^15^, Beck Anxiety Inventory (BECK-A)^16^ and Pennsylvania State Worry Questionnaire (PSWQ)^17^.

#### Secondary Outcomes

Assessment of conditions frequently associated with GAD, using validated instruments.

1. Depression will be measured using the Hamilton Depression Rating Scale (HAM-D)^18^ and/or the Beck Depression Inventory (BECK-D)^19^.
2. Insomnia will be assessed with a recognized questionnaire, such as the Pittsburgh Sleep Quality Index (PSQI)^20^.
3. Quality of life, assessed using a valid and reliable instrument, such as the World Health Organization Quality of Life (WHOQOL-BREF)^21^ and the 36-Item Short Form for Health Research (SF-36)^22^.

### Study designs

Due to the lack of studies in this area, we will include all randomized and non-randomized clinical trial, case-control, cohort and case series studies that evaluate the effectiveness of acupuncture and/or its related techniques for GAD.

Additionally, the following design information will be evaluated to confirm inclusion in the study:

1. studies that used valid and reliable instrument scales to assess quality of life;
2. studies that observed or used the acupuncture technique and/or its derivations to treat GAD;
3. in case of clinical trials, when the experimental group is used in addition to another active treatment, the study will be included;
4. reported outcomes must be about anxiety;
5. articles that present complete data regarding the primary and secondary outcomes adopted in this study;
6. complete texts published in English, Portuguese or Spanish.

### Search methods for identifying studies

The search will not be restricted by date or publication status.

#### Electronic searches

A search will be carried out in the following databases: EMBASE, LILACS, PubMed/MEDLINE, SCIELO, SCOPUS, CINAHL, Web of Science and Cochrane Library. The search will utilize Medical Subject Headings (MeSH) terms without restrictions on the publication period. Only articles published in English, Portuguese, or Spanish will be included. A comprehensive search strategy (Fig 1) will be applied.

**Fig 1.**
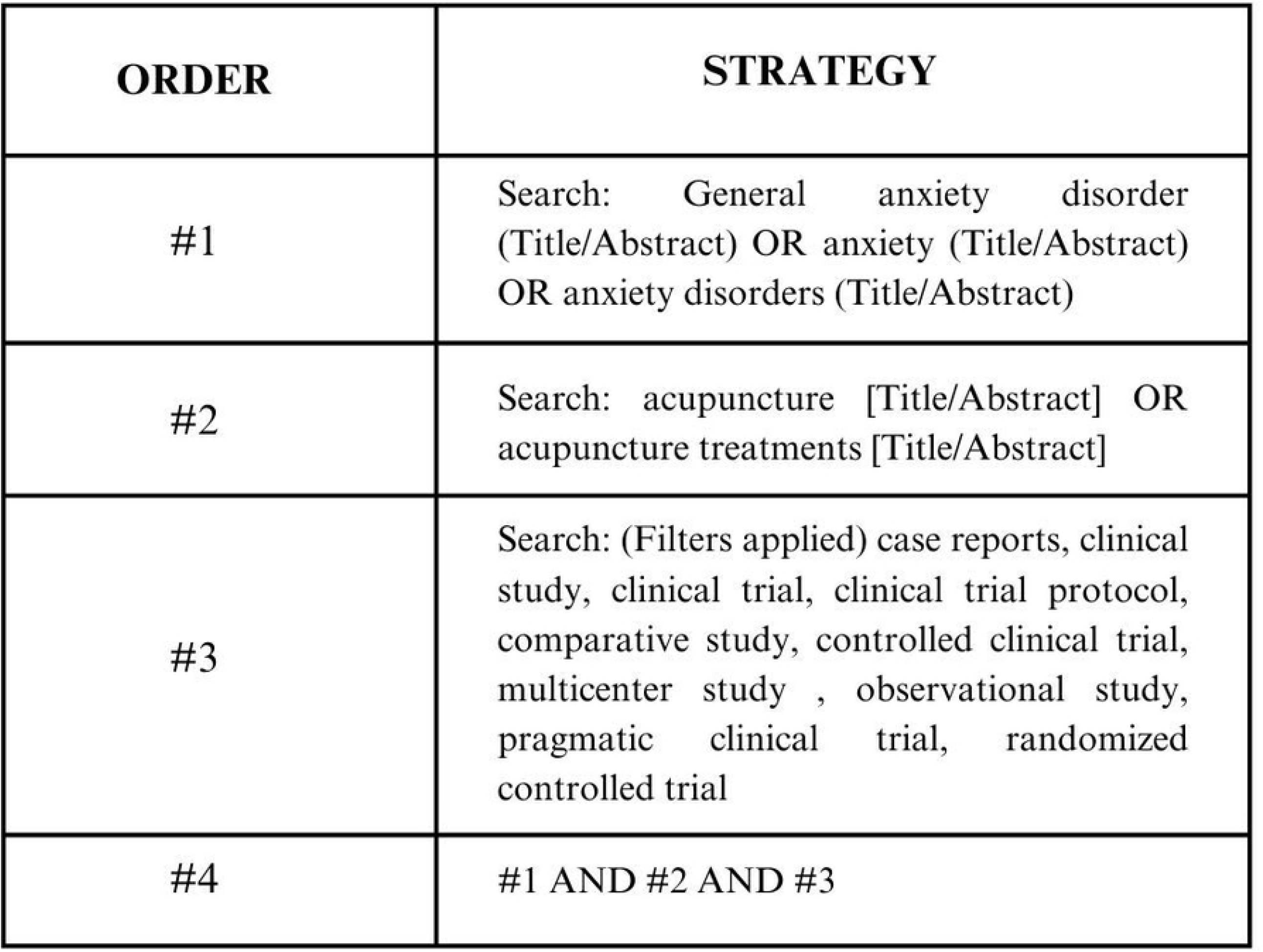
Search strategy for Pubmed.

#### Searching other resources

A search will be carried out on the topic in the gray literature in order to include as many studies carried out on the subject as possible.

#### Selection of studies

After searching the databases, all records obtained will be imported into the Ryyan reference management software^23^ to remove duplicates. Then, two independent reviewers (CCS and MAM) will screen the titles and abstracts to verify eligibility according to the inclusion criteria. Potentially eligible studies will be retrieved for full reading and any disagreement between reviewers will be resolved by consensus or by consultation with a third reviewer. The selection process will be documented using the PRISMA^24^ flowchart, presented in Fig 2.

**Fig 2.**
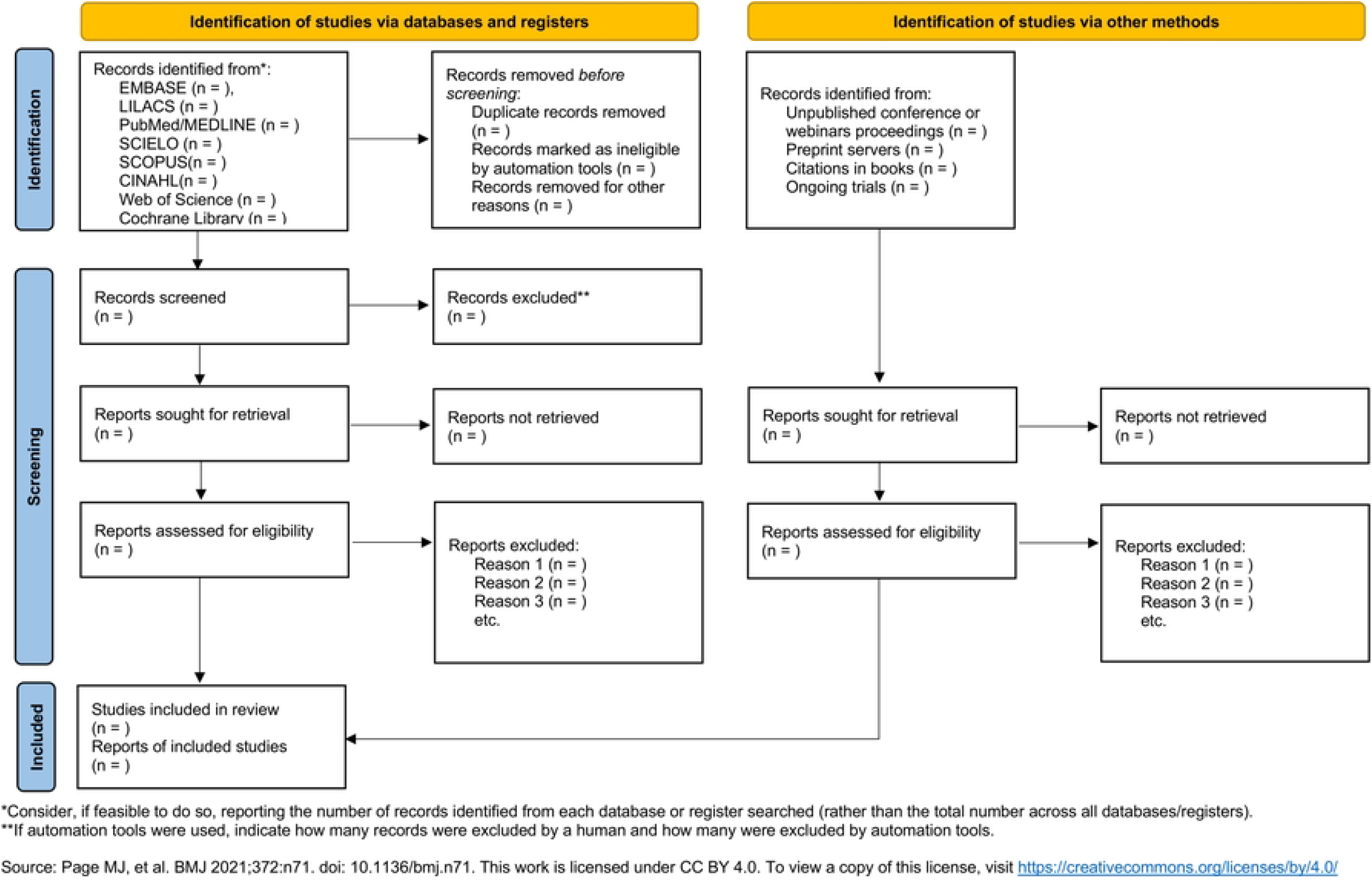
Search Flowchart.

### Data extraction and management

Two independent reviewers (CCS and MAM) will screen articles to include eligible studies by reading study titles and abstracts according to the inclusion and exclusion criteria. The full text of potentially eligible studies will be reviewed to confirm eligibility. When all included studies have been determined, the same two reviewers will perform data extraction in terms of author name, year of publication, study duration, types of intervention, outcomes (including unit of measurement and interpretation of scores). For clinical trials, information regarding groups, number of participants in each group, mean, SD), types of control, total number of participants, number of sessions, total number of participants in intervention groups, total number of participants in control groups will be added and psychometric instruments used. The values found will be evaluated using the Mean Difference (MD) and those with different metrics will be used the Standard Mean Difference (SMD). MD and SMD will be assessed with corresponding 95% confidence intervals (CI) for changes in pre- and post-intervention period measurements. Data extracted from clinical trials will be analyzed using Review Manager Software 8.1.1 (RevMan) (available at: http://revman.cochrane.org/). Any disagreements during data extraction will be resolved by consulting a third reviewer (SRRB).

### Assessment of risk of bias in included studies

We will use the Mixed Methods Appraisal Tool (MMAT) Version 2018^25^ and the Standards for Reporting Interventions in Clinical Trials of Acupuncture – STRICTA^26^ to assess the quality of the studies. In the evaluation phase, these tools will be applied by two reviewers, evaluating their quality and also reducing the assessment of bias. If there is controversy during the risk of bias assessment, it will be arbitrated by a third reviewer (SRRB).

### Handling Missing Data

If trial data are incomplete or unavailable, the corresponding author will be contacted via email to request additional information. If the data cannot be obtained or the author is unreachable, the study will be excluded from the analysis. A restricted analysis will be conducted using the available data, and the potential implications of missing information will be considered.

### Data synthesis

We will conduct a systematic review evaluating effect size measures in within- and between-interactions. The data will be synthesized depending on the measurements presented by the studies, following the Preferred Reporting Items for Systematic Reviews and Meta-analyses (PRISMA) statement^24^. Meta-analysis will only be considered if we find standardized measures and a comparable population between studies.

### Subgroup analysis

Subgroup analysis will be conducted to explore sources of heterogeneity when considerable heterogeneity is detected. If possible, subgroup analysis will be performed in terms of the following aspects: types of acupuncture (e.g., electroacupuncture, manual acupuncture, auriculotherapy, and moxibustion); types of control interventions; time points of primary outcome measurements; and age groups of patients.

### Patient and public involvement

Patients and/or the public were not involved in the design, or conduct, or reporting, or dissemination plans of this research.

### Ethics and dissemination

Ethical committee approval for this review is unnecessary since the data used will be extracted from pre-existing literature. The results will be submitted for publication in a peer-reviewed journal and presented at international academic conferences.

## Discussion

Generalized Anxiety Disorder (GAD) is a chronic mental health condition characterized by excessive and persistent worry, often accompanied by a range of physical symptoms. It significantly impacts patients’ quality of life, daily functioning, and overall well-being^1^. Conventional treatment typically involves pharmacological interventions alongside cognitive-behavioral therapy (CBT)^4^. However, long-term use of medications is associated with adverse effects, including dependence and withdrawal symptoms, highlighting the need for complementary and alternative therapeutic options^5^. Acupuncture, as a holistic intervention rooted in Traditional Chinese Medicine, has gained attention for its potential role in modulating the autonomic nervous system and neurochemical pathways implicated in anxiety regulation^9-12^. Previous systematic reviews have predominantly relied on Chinese clinical studies, which remain largely inaccessible in Western databases^7,8^. However, Cochrane reviewers have seldom incorporated Chinese sources into their search strategies^27^. Therefore, it is necessary to conduct a systematic review using accessible Western studies to fill this gap.

This review aims to assist clinicians in treating Generalized Anxiety Disorder (GAD) and provide valuable evidence for researchers. Patients may also benefit from new alternatives for their clinical management.

However, this review has limitations. Chinese databases will not be included due to their unavailability in the West. Additionally, there is likely to be significant heterogeneity among the studies, as non-randomized clinical trials may also be included. Despite these limitations, this systematic review is expected to enhance and expand the understanding of acupuncture as a treatment for GAD.

## Data Availability

No datasets were generated or analysed during the current study. All relevant data from this study will be made available upon study completion.

## Acknowledgements

I sincerely thank the professors and colleagues of the Master’s Program in Public Health at the Institute of Tropical Pathology and Public Health (IPTSP) – Federal University of Goiás (UFG), whose support and collaboration were essential throughout this process.

## Supporting information

**S1 Appendix. The PRISMA-P Checklist**.

